# Measles Virus Genomic Surveillance Gaps during a Nationwide Outbreak, Bangladesh, 2026

**DOI:** 10.64898/2026.06.24.26356456

**Authors:** Nehal Hasnain, Selim Farhad Shihab, Md. Asadul Islam, Md. Abdur Rahman

## Abstract

Bangladesh reported a nationwide measles outbreak in April 2026 involving over 19,000 suspected cases, despite high reported first-dose vaccine coverage (≥95%). We assessed whether publicly available molecular data could support epidemiologic interpretation of this resurgence and evaluated broader sequence sharing practices across South Asia. We analyzed public outbreak reports, WHO/UNICEF Estimates of National Immunization Coverage (WUENIC), PubMed indexed literature, and NCBI GenBank records from nine regional countries. Public sequence visibility across the region was highly uneven. While India and Pakistan associated records dominated the public dataset, only 32 Bangladesh origin records were retrieved, and notably, none were collected after 2019. The sole 2026 Bangladesh linked molecular record was a travel associated genotype B3 genome isolated in Australia (PZ189094.1). Its closest public N450 relative was a contemporaneous Pakistan sequence (2-nucleotide difference). The historical Bangladesh sequences were more distant, precluding robust phylogenetic inference regarding local viral persistence, cross-border importation, or transmission direction. Immunization data revealed a high regional baseline but highlighted subnational vulnerability and a significant pandemic era coverage collapse in neighboring Myanmar. The absence of timely, publicly available genomic data during the critical early months of the outbreak highlights a severe genomic surveillance gap. Public molecular records were historically sparse and insufficient to reconstruct outbreak transmission dynamics. To support elimination goals, establishing targeted sequencing pipelines, enforcing minimum metadata standards, and ensuring rapid public data deposition are urgently needed.

## Introduction

Measles is among the most transmissible vaccine preventable viral infections, and outbreaks can expand rapidly when immunity gaps accumulate. Bangladesh has made substantial progress toward measles control through routine immunization, supplementary immunization activities, and measles-rubella campaigns (1–4), but subnational immunity gaps, delayed second dose coverage, displaced populations, and post-disruption birth cohorts can still drive outbreaks. In April 2026, the World Health Organization (WHO) reported a nationwide increase in measles in Bangladesh affecting 58 of 64 districts across all 8 divisions. During March 15–April 14, 2026, Bangladesh reported 19,161 suspected cases, 2,897 laboratory confirmed cases, and 166 measles associated deaths; 79% of reported cases were among children <5 years of age (5). The outbreak created an urgent need not only for case detection and vaccination response but also for molecular context (6); the regional resurgence is part of a documented 2024–2026 global measles increase (7).

Molecular surveillance can strengthen measles outbreak investigation by linking epidemiologic patterns to viral genotype and sequence evidence. The 450-nt carboxyl-terminal region of the nucleoprotein gene (N450) is the WHO standardized window for genotype assignment and for comparing measles virus strains across outbreaks and countries. Dense, timely sequence data can help distinguish whether cases are compatible with local persistence, repeated importation, or multiple introductions; sparse public sequence data can prevent such interpretation even when outbreak reports document substantial transmission. South Asia has reported measles virus genotype diversity, including B3 and D8 circulation in India, Pakistan, Bangladesh, and neighboring countries (8–18). Public sequence visibility is uneven across the region. Countries with many public records may appear molecularly prominent even when this reflects deposition practices, whereas countries with few public records may have active transmission that remains poorly represented. For Bangladesh’s 2026 outbreak, this distinction is central. The absence of timely Bangladesh origin public sequences is the principal finding, not a minor data limitation.

The aim of this rapid public data analysis was to consolidate publicly available evidence on measles virus genotypes, regional molecular epidemiology, and genotype-based outbreak tracking in Bangladesh and South Asia, and to identify gaps in molecular surveillance and public sequence sharing that limited interpretation of the 2026 outbreak. We analyzed publicly available outbreak reports, literature, immunization estimates, and measles virus sequence records to assess whether public molecular evidence was sufficient to contextualize the outbreak and to identify genomic surveillance gaps relevant to outbreak response.

## Methods

### Data sources

This work is positioned as a Research article in the public data analysis sense. The contribution is the synthesis of existing public sources into an integrated regional evidence map of measles molecular surveillance, not the generation of new sequence or epidemiological data. We used 5 public information sources: WHO Disease Outbreak News for the 2026 Bangladesh outbreak (5), CDC global measles outbreak context (6), PubMed-indexed literature, NCBI Nucleotide/GenBank records for measles virus sequences, and WHO/UNICEF Estimates of National Immunization Coverage (WUENIC) for measles-containing-vaccine (MCV) coverage. All searches were conducted on May 21, 2026. A no-date-filter requery on June 16, 2026 returned 169, 97, and 30 records for the three searches, indicating 3 additional records for Search 1 and 4 for Search 2 had been indexed in PubMed after May 21; the four new records are listed in the Appendix. None of these post-May-21 records changed the regional literature-density interpretation. This study used public data only; ethics review was not required.

### Search strategy and evidence selection

This study was a rapid public data analysis and evidence map, not a formal systematic review or meta-analysis. Although not a formal systematic review, we adapted principles from the PRISMA-ScR (Scoping Reviews) guidelines to structure our literature search. PubMed searches identified peer-reviewed evidence on measles outbreaks, molecular epidemiology, genotype detection, vaccination context, and surveillance in Bangladesh and the broader South Asian region (the 8 South Asian Association for Regional Cooperation [SAARC] countries: Afghanistan, Bangladesh, Bhutan, India, Maldives, Nepal, Pakistan, Sri Lanka plus Myanmar). The searches prioritized specificity by using Title/Abstract terms; this strategy reduced irrelevant records but may have missed relevant reports that did not use the selected terms in title or abstract. The 3 PubMed searches were: (1) measles or measles virus AND any SAARC country (Afghanistan, Bangladesh, Bhutan, India, Maldives, Nepal, Pakistan, Sri Lanka) or Myanmar AND genotype, molecular, sequencing, phylogenetic, or outbreak; (2) measles or measles virus AND Bangladesh AND outbreak, resurgence, surveillance, or vaccination; and (3) measles virus AND genotype AND any SAARC country or Myanmar. Records were considered relevant if they described measles outbreak context, genotype or sequence data, molecular detection, phylogenetic analysis, vaccination context, or surveillance information relevant to Bangladesh or the broader South Asian region. We used the literature to contextualize public molecular evidence rather than to estimate pooled effects.

Yields on the May 21, 2026 search date were 166 records for Search 1, 93 records for Search 2, and 30 records for Search 3. Per-country counts for Search 1 were: India 111, Pakistan 21, Bangladesh 17, Afghanistan 8, Nepal 8, Myanmar 8, Sri Lanka 7, Bhutan 1, Maldives 0 (sum 181; the OR-merged Search 1 total is 166 because some records are tagged to multiple countries). The post May 21 re-query (June 16, 2026) returned 169, 97, and 30 records; the 3 additional Search 1 records (a Pakistan 2022–2023 outbreak study, a Bangladesh resurgence commentary, a 2026 Bangladesh news article) and 4 additional Search 2 records (the same three plus a Bangladesh SSPE case series) are listed in the Appendix and did not change the regional literature density interpretation.

### GenBank sequence retrieval and curation

NCBI Nucleotide was queried using “txid11234[Organism:exp]” combined with country terms for the 8 SAARC countries plus Myanmar. Records were retrieved as GenBank XML through NCBI E-utilities and de-duplicated by unique accession number. For each record we parsed accession number, raw definition, organism, geographic origin (geo_loc_name), strain and isolate names, collection date, and genotype label where present. Two complementary denominators were used and are reported explicitly: country-term search hits, which describe the publicly visible sequence landscape an investigator encounters, and (ii) geographic origin parsed from each unique record, used to summarize genotype composition. Country-associated counts were interpreted as indicators of public sequence visibility, not as national incidence, prevalence, or reservoir size.

We distinguished locally collected country-associated records from travel-associated records when metadata indicated collection outside South Asia but travel or importation linked to a South Asian country. The 2026 accession PZ189094.1 was classified as travel-associated with Bangladesh because the record was collected in Queensland, Australia, and its source qualifiers indicated travel in Bangladesh (19).

### Genotype and geographic classification

Genotype labels were extracted from standardized measles strain names and bracketed labels (e.g., [B3], [D8], [D9]) when available. Because GenBank country-term searches can retrieve records through isolate names, source qualifiers, submitter fields, publication text, or travel-associated metadata, we did not treat country search counts as curated national totals; we used them to describe the publicly visible sequence landscape available during outbreak interpretation. Genotypes were taken from submitter labels rather than re-called against WHO reference strains or WHO MeaNS, and no inter-rater validation of geographic origin was performed; this is noted as a limitation.

### Sequence comparison

For accession-level comparison we prioritized homologous N450 regions. The full comparison set in Figure 2 comprised 22 records (19 Bangladesh B3 + 2 Meghalaya B3 + 1 West Bengal B3); the 12-taxon focused alignment in Appendix Table 3 is a representative subset for table legibility and does not alter the central identity result. Sequences were aligned with MAFFT (`--auto`) and percent nucleotide identity was calculated across 450-nt N-gene windows excluding pairwise gap sites and ambiguous bases; the 22-taxon Figure 2 alignment used 450 comparable columns and the 12-taxon Appendix Table 3 alignment used 447, with the small numeric difference (98.889% vs 98.881% in the same pairwise comparison) reflecting only the column count (Appendix). An exploratory UPGMA dendrogram, including a contemporaneous 2026 Pakistan B3 comparator, is provided as an exploratory aid only (Appendix Figure 1). We compared the Queensland travel-associated B3 sequence PZ189094.1 with historical Bangladesh B3 records (2014-2019), northeastern Indian Meghalaya B3 records (KX350060.1, KX350059.1) and West Bengal (Basirhat) B3 record (MW735989.1). Sequence similarity was interpreted as evidence of relatedness within a regional B3 background, not as proof of direct ancestry, origin, transmission direction, or local persistence. Because most regional records are 450-nt N-gene fragments rather than whole genomes, no full-genome phylogeny or transmission-chain reconstruction was attempted.

### Immunization data and analysis

WUENIC MCV1 and MCV2 estimates for 2000–2024 were used as epidemiologic context(20). For each of the 9 countries with WUENIC series, we extracted the 2024 estimate, the 25-year mean and range, pre-pandemic (2010–2019) versus pandemic-era (2020–2024) means, and the MCV1–MCV2 gap. 2025 administrative coverage, where reported, was compared with 2024 WUENIC. Coverage estimates were used as context and were not used to explain genotype distributions or GenBank record counts.

## Results

### Bangladesh 2026 outbreak context

WHO reported that Bangladesh notified a nationwide increase in measles on April 4, 2026, affecting 58 of 64 districts across all 8 divisions. During March 15–April 14, 2026, 19,161 suspected cases, 2,897 laboratory confirmed cases, and 166 measles associated deaths were reported; 79% of cases were among children <5 years of age (5). WHO also described a targeted measles-rubella vaccination campaign beginning April 5, 2026, with strengthened surveillance and epidemiologic analysis. These data documented a large national outbreak but did not provide public accession level viral sequence evidence from Bangladesh collected 2026 cases.

### Published molecular evidence from Bangladesh and South Asia

PubMed searches identified regional literature describing measles virus genotype circulation, molecular detection, and outbreak investigation. Among the 166 records returned by Search 1, India accounted for 111 (66.9%), Pakistan 21, Bangladesh 17, and Afghanistan, Nepal, and Myanmar 8 each; Sri Lanka contributed 7, Bhutan 1, and Maldives 0. (Per-country counts include records tagged to multiple countries; the May 21 Search 1 cohort is 166, but the sum of per-country counts is 181. The country-specific numbers reflect the OR-merged regional literature landscape.) The distribution was strongly skewed toward India, paralleling the GenBank public sequence visibility pattern. India and Pakistan had multiple studies reporting genotype diversity, genotype B3 detection, D8 circulation, lineage patterns, and laboratory network expansion (9–18, 21, 22). Bangladesh specific molecular literature was more limited in the retrieved corpus, although one recent cross-sectional study reported molecular detection and genetic diversity of measles and parainfluenza viruses circulating in humans in Bangladesh (8). Historical Bangladesh labelled GenBank records document genotype B3 and D8 sequences, including Cox’s Bazar B3, Mymensingh D8, and Chandpur B3 accessions (23, 24, 25); Nepal examples included D8 records such as KX455932.1 (26). Notably, the broader SAARC plus Myanmar search did not retrieve additional molecular epidemiology studies for Bhutan, Maldives, or Sri Lanka beyond what the original narrower search returned, reinforcing the regional literature-density asymmetry.

### Regional public sequence visibility

Country-term searches across the 9 country terms returned 4,151 country-associated hits, which de-duplicated to 4,109 unique accession-level records. By search hits, India accounted for 2,345 records, Pakistan 1,645, Nepal 44, Afghanistan 42, Bangladesh 34, Myanmar 28, Sri Lanka 10, Bhutan 2, and Maldives 1 (Table 1). Among the 4,109 unique records geolocated to country of origin, genotype composition was strongly country specific (Appendix Table 2). India origin records were predominantly D8 (1,903/2,227; 85.5%), Pakistan origin predominantly B3 (1,535/1,592; 96.4%), Afghanistan origin B3 (33/40; 82.5%), and Myanmar origin D9 (27/28; 96.4%); the 32 Bangladesh origin records comprised B3 (19; 59.4%) and D8 (7; 21.9%), with 6 unlabelled. These counts should be read as public sequence visibility, not transmission burden. The key result for Bangladesh was the small number and historical nature of publicly visible Bangladesh associated records during a major 2026 outbreak.

**Table 1.** Public data sources and outputs used for the rapid evidence map, Bangladesh and South Asia, 2026.

| Source | Search focus | Search date | Output used | Interpretation |
| --- | --- | --- | --- | --- |
| WHO Disease Outbreak News | Bangladesh 2026 measles outbreak | May 21, 2026 | 1 outbreak report | Current outbreak statistics and response context |
| CDC global measles outbreak page | Global outbreak and travel context | May 21, 2026 | 1 web source | Cross-border measles risk context |
| PubMed | Bangladesh and SAARC+Myanmar measles outbreak, genotype, molecular surveillance | May 21, 2026 | 3 searches: 166, 93, 30 records | Peer-reviewed context; not an exhaustive systematic review |
| NCBI Nucleotide | txid11234[Organism:exp] plus 9 regional country terms | May 21, 2026 | 4,151 hits (4,109 unique records) | Public sequence visibility indicator (frozen at May 21, 2026) |
| WUENIC | MCV1/MCV2 coverage estimates | 2000–2024 | National coverage trends | Epidemiologic context only |

### Bangladesh public sequence gap

We identified 34 Bangladesh associated GenBank records through the search strategy; 32 geolocated to Bangladesh, and all were from 2014 to 2019. No Bangladesh origin record carried a collection date later than 2019 (Figure 1). Across the entire post 2020 period, the de-duplicated dataset contained 1,251 Pakistan origin and 177 India origin records but none of Bangladesh origin. The only Bangladesh linked 2026 molecular record was PZ189094.1, collected in Queensland, Australia, with source qualifiers indicating travel in Bangladesh (19). Public epidemiologic data documented nationwide transmission, but public molecular data did not provide enough Bangladesh origin sequences to reconstruct outbreak spread, compare districts, or distinguish plausible introduction scenarios.

**Figure 1.**
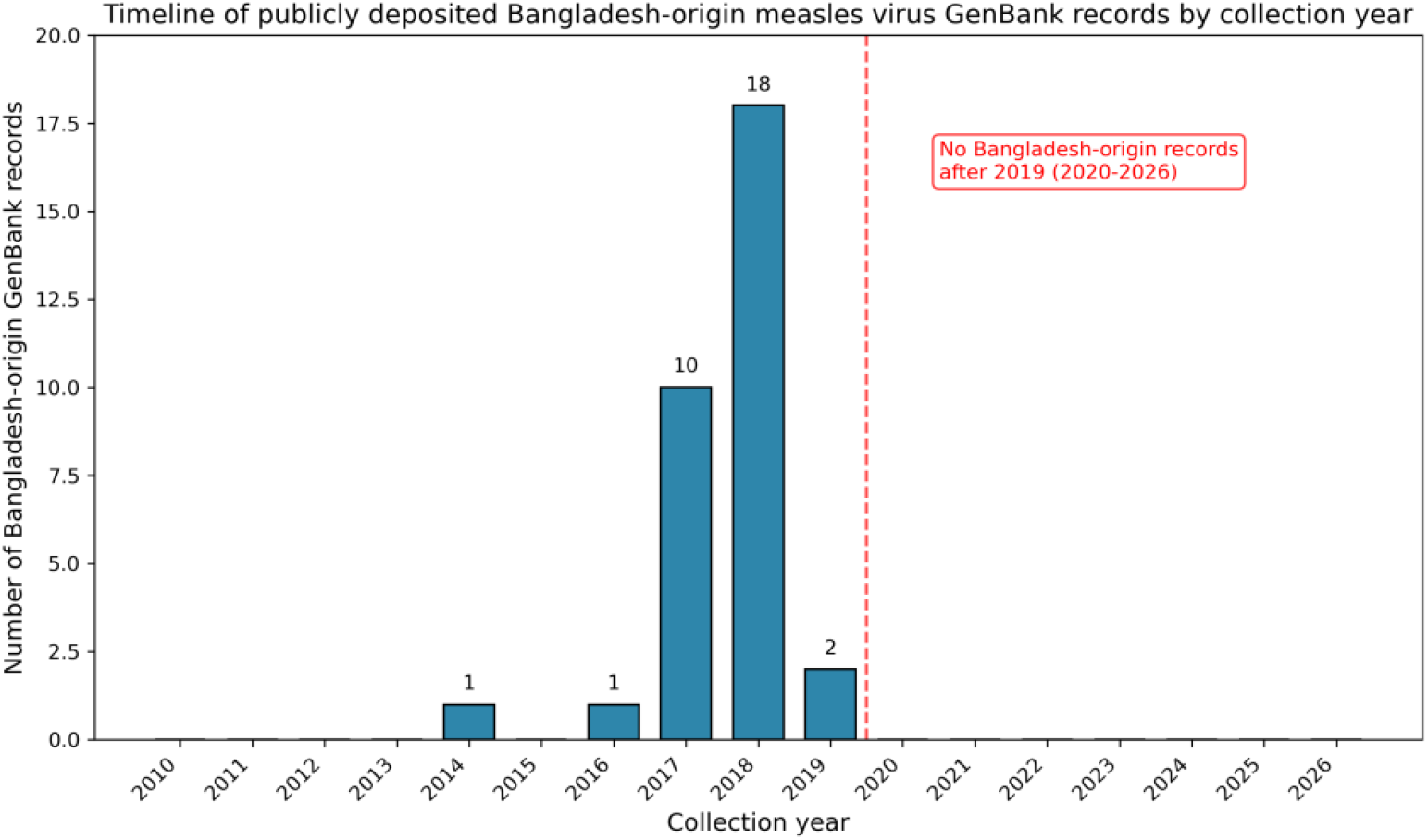
Timeline of publicly deposited Bangladesh-origin measles virus GenBank records by collection year.

### Travel-associated PZ189094.1 and N450 similarity

PZ189094.1 is a 2026 genotype B3 complete-genome record (strain MVs/Queensland.AUS/02.26[B3]) collected in Queensland, Australia, and linked by source metadata to travel in Bangladesh (19). In the homologous 450-nt N-gene window, PZ189094.1 showed 98.889% nucleotide identity (445/450 matches; 5 differences) with 10 historical Bangladesh B3 accessions (KX671298.1, MK161512.1, MK161514.1, MK161516.1, MK183782.1, MK183783.1, MK183784.1, MK628286.1, MK628292.1, MK628293.1). The same 98.889% identity was observed with the West Bengal B3 accession MW735989.1 and the Meghalaya B3 accessions KX350060.1 and KX350059.1 (27, 28), which were identical to each other across N450. Furthermore, 9 Bangladesh B3 records showed 98.667% identity (444/450) (Figure 2). The same 5-nt distance was therefore observed for both the historical Bangladesh B3 records, the Basirhat B3 record in southeastern West Bengal near Bangladesh’s western border, and the Meghalaya B3 records in northern India bordering Bangladesh’s northeastern side. PZ189094.1 represents at least one lineage (B3) circulating in Bangladesh during the 2026 period but does not exclude concurrent D8 or other genotype circulation, which cannot be evaluated from the public data.

**Figure 2.**
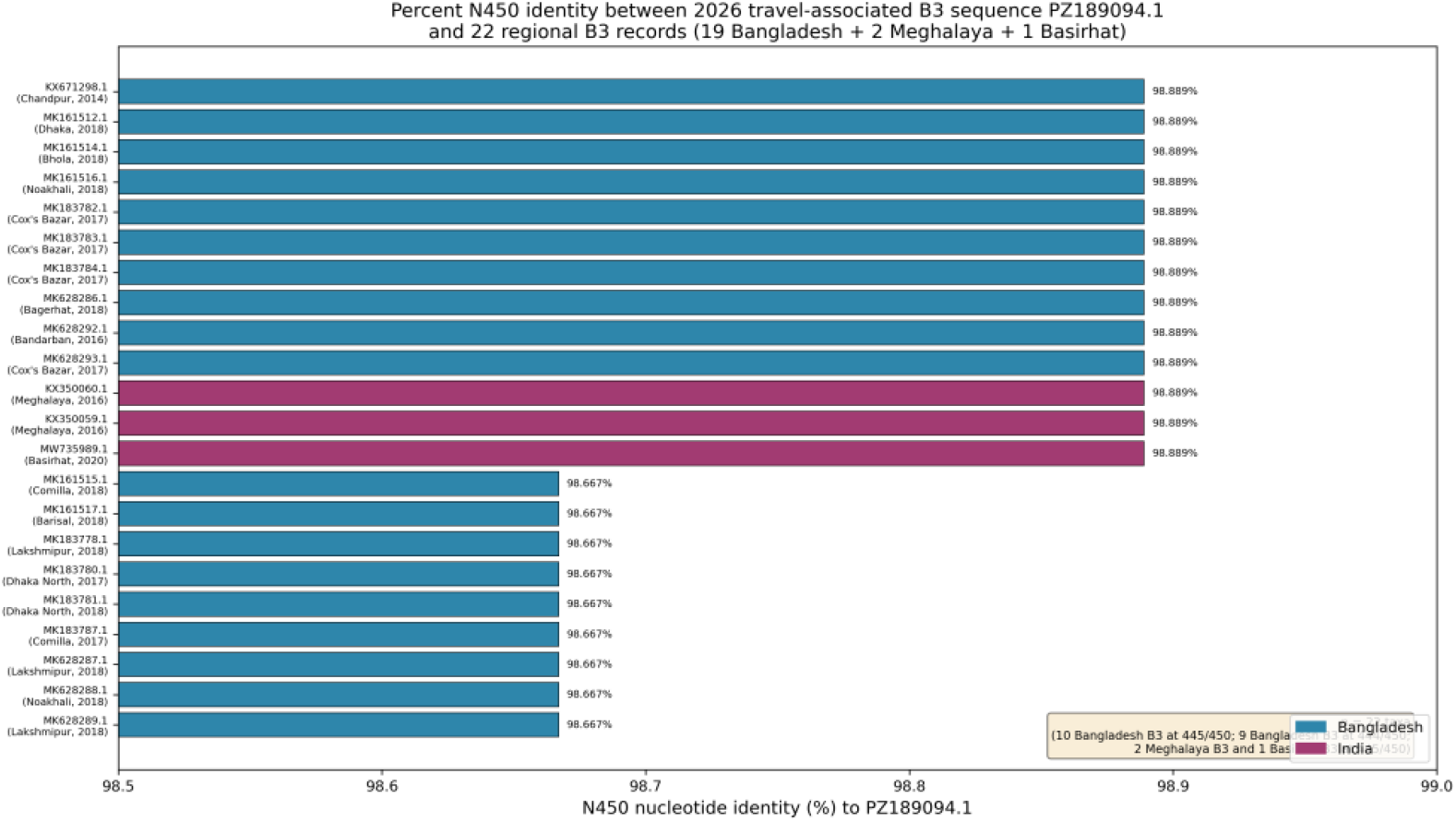
Horizontal bar chart of N450 (450-nt nucleoprotein-gene window) percent nucleotide identity between the 2026 travel-associated genotype B3 sequence PZ189094.1 and 22 regional B3 records (19 Bangladesh B3 + 2 Meghalaya B3 + 1 Basirhat B3). Note: *Bars are sorted by descending identity and color-coded by country of origin. PZ189094*.*1 is 98*.*889% identical (445/450) to both the northeastern Indian (Meghalaya) sequences, the Basirhat B3 record, and 10 historical Bangladesh B3 accessions; 9 further Bangladesh B3 records show 98*.*667% identity (444/450). The equal similarity to Indian and Bangladeshi records indicates a shared regional B3 background and precludes inference of geographic origin or transmission direction*.

A 12-taxon focused MAFFT alignment yielded the same pairwise result at 98.881% over 447 comparable columns (Appendix Table 3); the small numeric difference reflects only the column count and not biological difference.

Critically, the northern (West Bengal) and northeastern Indian (Meghalaya) sequences were equally similar to PZ189094.1 as the Bangladesh sequences (the identical 98.889% value), and the single closest public N450 relative was not a Bangladesh sequence but a contemporaneous 2026 Pakistan B3 record (PZ106574.1, Bajaur(29)), which differed from PZ189094.1 by only 2 nucleotides versus 5 for the historical Bangladesh, West Bengal and Meghalaya records (Appendix Table 3; Appendix Figure 1). This pattern indicates that the 2026 travel associated sequence belongs to a regional B3 background shared across Bangladesh, northeastern India, and Pakistan, but it cannot localize the lineage to Bangladesh, identify a direct ancestor, establish transmission direction, or demonstrate that a hidden endemic lineage persisted between 2020 and 2026. A difference of only a few nucleotides in a 450-nt fragment provides limited phylogenetic resolution and supports relatedness only, not ancestry; the nearest public relative being Pakistani rather than Bangladeshi underscores that public data may not identify the geographic source.

### Immunization context

WUENIC estimates for 2000–2024 showed marked regional heterogeneity in MCV coverage (Table 2). In 2024, MCV1 ranged from 55% in Afghanistan and 71% in Myanmar to 96–99% in Bangladesh, Bhutan, India, Maldives, Nepal, and Sri Lanka; MCV2 ranged from 44% in Afghanistan to 92–99% in the high-coverage group. The MCV1–MCV2 gap was narrow (0–5 percentage points) in most countries but reached 11 percentage points in Afghanistan.

**Table 2.**
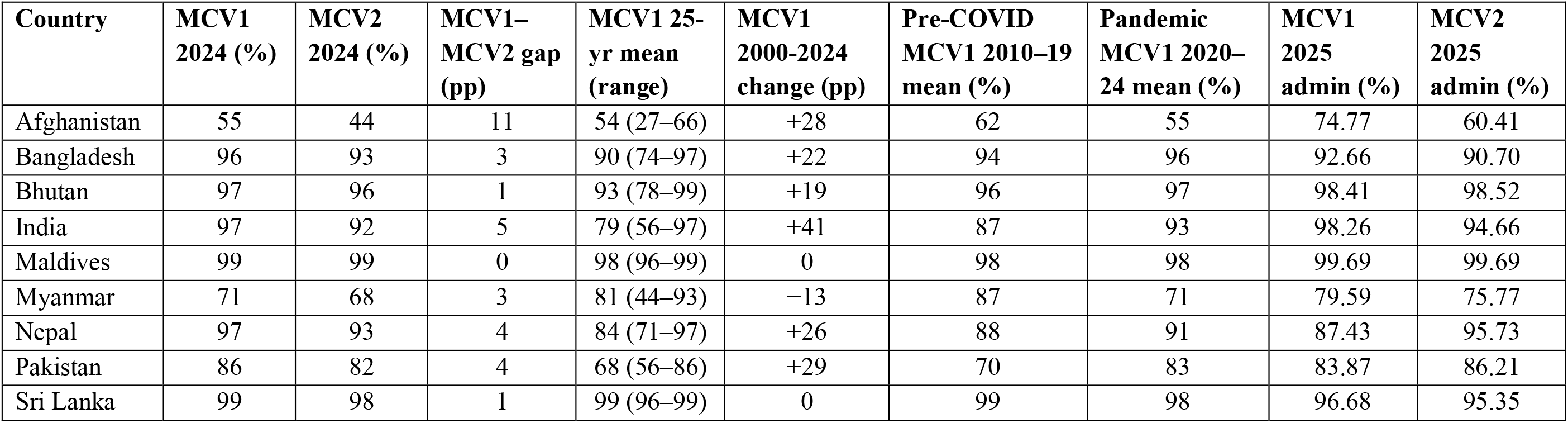
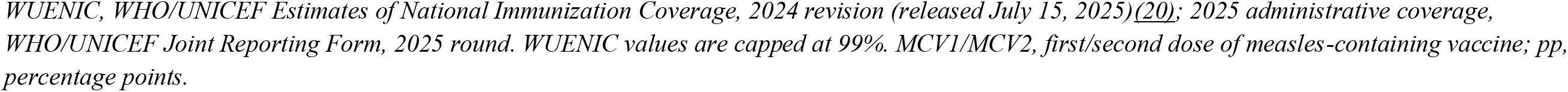
Measles-containing-vaccine coverage in South Asia: 25-year WUENIC trends (2000–2024) and 2025 administrative coverage.

| Country | MCV1 2024 (%) | MCV2 2024 (%) | MCV1–MCV2 gap (pp) | MCV1 25-yr mean (range) | MCV1 2000–2024 change (pp) | Pre-COVID MCV1 2010–19 mean (%) | Pandemic MCV1 2020–24 mean (%) | MCV1 2025 admin (%) | MCV2 2025 admin (%) |
| --- | --- | --- | --- | --- | --- | --- | --- | --- | --- |
| Afghanistan | 55 | 44 | 11 | 54 (27–66) | +28 | 62 | 55 | 74.77 | 60.41 |
| Bangladesh | 96 | 93 | 3 | 90 (74–97) | +22 | 94 | 96 | 92.66 | 90.70 |
| Bhutan | 97 | 96 | 1 | 93 (78–99) | +19 | 96 | 97 | 98.41 | 98.52 |
| India | 97 | 92 | 5 | 79 (56–97) | +41 | 87 | 93 | 98.26 | 94.66 |
| Maldives | 99 | 99 | 0 | 98 (96–99) | 0 | 98 | 98 | 99.69 | 99.69 |
| Myanmar | 71 | 68 | 3 | 81 (44–93) | –13 | 87 | 71 | 79.59 | 75.77 |
| Nepal | 97 | 93 | 4 | 84 (71–97) | +26 | 88 | 91 | 87.43 | 95.73 |
| Pakistan | 86 | 82 | 4 | 68 (56–86) | +29 | 70 | 83 | 83.87 | 86.21 |
| Sri Lanka | 99 | 98 | 1 | 99 (96–99) | 0 | 99 | 98 | 96.68 | 95.35 |

WUENIC, WHO/UNICEF Estimates of National Immunization Coverage, 2024 revision (released July 15, 2025)(20); 2025 administrative coverage, WHO/UNICEF Joint Reporting Form, 2025 round. WUENIC values are capped at 99%. MCV1/MCV2, first/second dose of measles-containing vaccine; pp, percentage points.

**Table 3.** Key sequence records relevant to Bangladesh 2026 molecular context.

| Accession | Country/link | Collection context | Genotype | Relevance |
| --- | --- | --- | --- | --- |
| PZ189094.1 | Queensland, Australia;<br>travel in Bangladesh | 2026 travel-associated<br>complete genome | B3 | Rare public molecular signal linked to Bangladesh during outbreak period; not evidence of community circulation |
| KX671298.1 | Chandpur, Bangladesh | 2014 historical<br>Bangladesh record | B3 | 98.889% N450 identity with PZ189094.1 |
| MK628293.1 | Cox's Bazar, Bangladesh | 2017 historical<br>Bangladesh record | B3 | 98.889% N450 identity with PZ189094.1 |
| MW735989.1 | Basirhat, West Bengal,<br>India | 2020 North India record | B3 | 98.889% N450 identity with PZ189094.1 |
| KX350060.1 | East Khasi Hills,<br>Meghalaya, India | 2016 northeastern India<br>record | B3 | 98.889% N450 identity with PZ189094.1 |
| KX350059.1 | East Khasi Hills,<br>Meghalaya, India | 2016 northeastern India<br>record | B3 | 98.889% N450 identity with PZ189094.1; identical to KX350060.1 |
| PZ106574.1 | Bajaur, Pakistan | 2026 regional<br>comparator | B3 | Closest public N450 match to PZ189094.1 (2-nt difference); contemporaneous regional B3, not evidence of Bangladesh origin; geographically closer to Afghanistan than to Bangladesh (26) |
| MK628291.1 | Mymensingh, Bangladesh | 2018 historical<br>Bangladesh record | D8 | Demonstrates historical Bangladesh D8 visibility |

Twenty five-year trends diverged sharply (Table 2). India recorded the largest absolute MCV1 gain (+41 percentage points, 56% to 97%), followed by Pakistan (+29), Afghanistan (+28), and Nepal (+26%). Sri Lanka and Maldives remained flat at 99%. Myanmar was the regional outlier and the only country with a net MCV1 decline (−13 percentage points, 84% to 71%). The pandemic-era (2020–2024) MCV1 mean was lower than the pre-pandemic (2010–2019) mean in Afghanistan (−6.7 pp), Myanmar (−16.1 pp), and Sri Lanka (−1.0 pp), and higher elsewhere.

Despite high national MCV1 coverage in Bangladesh (96% in 2024; 25-year mean 90%), the 2026 nationwide outbreak occurred. National averages masked subnational heterogeneity documented in prior studies, including zero-dose clustering, persistent MCV2 lag, and pandemic-era service disruption (1–4, 30, 31, 32). 2025 administrative coverage, the first data released after the 2026 outbreak, was 92.66% MCV1 / 90.70% MCV2 in Bangladesh, 83.87% / 86.21% in Pakistan, and 79.59% / 75.77% in Myanmar. Administrative values were within 2–4 percentage points of 2024 WUENIC for most countries, except Myanmar, where administrative coverage was 8–9 points higher, suggesting possible over-reporting.

## Discussion

PZ189094.1 matters because it provides a rare public molecular signal linked to Bangladesh travel during the outbreak period. Its N450 region was highly similar to historical Bangladesh B3 records, to B3 sequences from Basirhat, West Bengal and East Khasi Hills, Meghalaya, indicating membership in a regional B3 background. However, the West Bengal, Meghalaya and Bangladesh sequences were equally similar to PZ189094.1, and its single closest public N450 relative was a contemporaneous 2026 Pakistan B3 sequence rather than a Bangladesh one. N450 similarity across a small set of public records cannot establish geographic origin, direct ancestry, or transmission direction, and cannot demonstrate continuous local persistence through the 2019–2026 sequence gap. Public data cannot distinguish local persistence, repeated introduction, or broader regional circulation.

The uneven GenBank landscape illustrates public sequence visibility bias. India and Pakistan dominated the retrieved regional dataset, whereas Bangladesh had few records and none after 2019. GenBank is a map of what was sequenced and shared, not of true transmission intensity; this limitation has been highlighted in the outbreak data-sharing literature (33). For outbreak response, the distinction is critical because sparse public deposits can distort risk perception. The deposition step appears to be at least one limiting step. Three explanations for the Bangladesh gap are each consistent with the public data: no domestic sequencing, internal sequencing without GenBank deposition, or deposition in a non-public database, and the data alone cannot adjudicate among them. PZ189094.1 was sequenced by a WHO Global Specialized Laboratory network member (Queensland Health, WHO Western Pacific Region reference laboratory), showing that regional GSL/LabNet capacity exists but is unevenly activated (34, 35). Investment in regional GenBank deposition workflows, minimum metadata standards, and cross-border data harmonization would convert the existing but unevenly shared sequence resource into an interpretable surveillance network. Sequencing and GenBank submission costs constrain LMIC laboratories and GAVI eligible support is a recognized policy lever (35).

Bangladesh’s high national MCV1 coverage does not rule out a nationwide outbreak. Measles can spread when local immunity gaps accumulate, when second-dose coverage lags, when zero-dose children cluster geographically or socially, when displaced populations remain vulnerable, or when pandemic-era disruptions leave susceptible cohorts. First-dose coverage alone is insufficient to interrupt transmission of a pathogen with very high basic reproduction number, and sustained high two-dose coverage is required (36). Persistent MCV1–MCV2 gaps in parts of the region, documented zero-dose clustering in urban and hard-to-reach populations, and vulnerability among forcibly displaced Rohingya populations in Cox’s Bazar are plausible structural contributors to susceptibility (30, 31), and COVID-19 era disruption of routine immunization is a documented regional stressor (32). The age distribution reported by WHO, with most cases among children <5 years of age, is consistent with the importance of recent immunity gaps (5). However, vaccination data cannot identify viral introduction routes or lineage persistence without linked molecular data.

The regional vaccination picture sharpens rather than weakens the surveillance gap finding. South Asia is no longer uniformly measles susceptible, as Sri Lanka, Bhutan, Bangladesh, Maldives, Nepal, and now India reported MCV1 above 95% and MCV2 above 90% in 2024. Bangladesh’s 2026 outbreak, in a country with sustained 90%+ MCV1 coverage, indicates that national elimination thresholds are not sufficient when immunity is geographically or demographically clustered. Myanmar is the regional exception, with 2024 MCV1 of 71% and 2021 coverage as low as 44%, a collapse attributable to disruption of routine immunization services and cold-chain access following the February 2021 military coup and continuing post-coup humanitarian and health-system constraints. The juxtaposition of high coverage without sequences (Bangladesh) and lower coverage with sporadic sequences (Afghanistan, Myanmar) defines the actionable gap because routine sequencing capacity and standardized public deposition are both needed, and the two cannot substitute for each other.

A practical surveillance response does not require sequencing every case. Bangladesh could prioritize targeted sequencing of early cases from district clusters, border district cases, airport or travel associated cases, Cox’s Bazar or Rohingya linked clusters, severe or fatal cases, and vaccine breakthrough cases. Minimum public metadata should include district, collection date, age, vaccination status, travel history, outbreak identifier, specimen type, RT-PCR result, sequence region, genotype, and accession number. Rapid N450 deposition, even before full genome capacity is universal, would substantially improve outbreak interpretation.

In conclusion, Bangladesh’s 2026 measles outbreak illustrates the difference between outbreak detection and molecular outbreak interpretation. Public data documented a nationwide outbreak, but public genomic data were too sparse to determine whether transmission reflected local persistence, repeated importation, or multiple introductions. PZ189094.1 is a valuable molecular signal that also highlights the absence of timely domestic public sequencing. Targeted sequencing, standardized metadata, and rapid public deposition should become part of measles outbreak response in Bangladesh and South Asia. This study has limitations. PubMed searches prioritized Title/Abstract specificity and may have missed non-indexed, non-English, or regional reports. GenBank country-term searches are non-representative and can retrieve records through multiple metadata fields, and genotypes were taken from submitter labels rather than recalled against WHO reference strains or WHO MeaNS. N450 comparison has limited resolution compared with whole-genome sequencing, and only a single travel-associated 2026 sequence was available, which cannot represent the Bangladesh outbreak. WUENIC national coverage estimates cannot explain subnational outbreak dynamics. The 2026 outbreak was still evolving when the search was conducted.

## Data Availability

All sequence records are available from NCBI GenBank under the accession numbers cited in the manuscript. WUENIC estimates are available from https://immunizationdata.who.int/. CDC outbreak data are available from https://www.cdc.gov/global-measles-vaccination/data-research/global-measles-outbreaks/index.html. All analysis scripts and the de-duplicated regional GenBank dataset are deposited at Zenodo (https://doi.org/10.5281/zenodo.20827758).

## Data and code availability

All analyses were conducted using publicly available data. The de-duplicated regional GenBank dataset, the N450 sequence alignments, pairwise-identity outputs, WUENIC extraction data, and all Python scripts used for data retrieval, sequence comparison, and figure generation are publicly available on Zenodo (https://doi.org/10.5281/zenodo.20827758). All findings are fully reproducible from this repository.

## Author contributions

NH designed the study. NH and SFS performed the analyses and drafted the manuscript. MAR and MAI all provided critical input on the analysis, as well as on the drafted manuscript.

