## Supplementary Appendix for "Measles Virus Genomic Surveillance Gaps during a Nationwide Outbreak, Bangladesh, 2026"

**Supplementary methods, data, and figures for:** Measles Virus Genomic Surveillance Gaps during a Nationwide Outbreak, Bangladesh, 2026

**Nehal Hasnain**, Department of Microbiology and Parasitology, Sher-e-Bangla Agricultural University (SAU), Dhaka, Bangladesh..

This Appendix provides the search strategies, the regional GenBank dataset summary, the full genotype-composition and pairwise N450 identity tables, an exploratory phylogenetic figure, the immunization-data methods, and the reproducibility resources supporting the main article. The main article stands alone; this material is provided for transparency and reproducibility. All analyses used public data accessed on May 21, 2026; sequence/identity re-checks on June 15, 2026.

#### Appendix Section 1. PubMed search strategy

PubMed was searched on May 21, 2026, using 3 specificity-focused queries restricted to the Title/Abstract fields, with country terms covering the 8 South Asian Association for Regional Cooperation (SAARC) countries like Afghanistan, Bangladesh, Bhutan, India, Maldives, Nepal, Pakistan, Sri Lanka plus Myanmar. These searches were used to identify peer-reviewed outbreak, genotype, molecular-surveillance, and vaccination-context evidence; they were not intended to constitute a formal PRISMA-ScR systematic review, and yields are reported as the counts returned on the search date.

**Operational yields (May 21, 2026):**

`(measles[Title/Abstract] OR "measles virus"[Title/Abstract]) AND (Afghanistan[Title/Abstract] OR Bangladesh[Title/Abstract] OR Bhutan[Title/Abstract] OR India[Title/Abstract] OR Maldives[Title/Abstract] OR Nepal[Title/Abstract] OR Pakistan[Title/Abstract] OR "Sri Lanka"[Title/Abstract] OR Myanmar[Title/Abstract]) AND (genotype[Title/Abstract] OR molecular[Title/Abstract] OR sequencing[Title/Abstract] OR phylogenetic[Title/Abstract] OR outbreak[Title/Abstract])` — 166 records.

`(measles[Title/Abstract] OR "measles virus"[Title/Abstract]) AND Bangladesh[Title/Abstract] AND (outbreak[Title/Abstract] OR resurgence[Title/Abstract] OR surveillance[Title/Abstract] OR vaccination[Title/Abstract])` — 93 records.

`"measles virus"[Title/Abstract] AND genotype[Title/Abstract] AND (Afghanistan[Title/Abstract] OR Bangladesh[Title/Abstract] OR Bhutan[Title/Abstract] OR India[Title/Abstract] OR Maldives[Title/Abstract] OR Nepal[Title/Abstract] OR Pakistan[Title/Abstract] OR "Sri Lanka"[Title/Abstract] OR Myanmar[Title/Abstract])` — 30 records.

**Per-country distribution (Search 1, May 21, 2026):**

| Country | Records |
|---|---|
| India | 111 |
| Pakistan | 21 |
| Bangladesh | 17 |
| Afghanistan | 8 |
| Nepal | 8 |
| Myanmar | 8 |
| Sri Lanka | 7 |
| Bhutan | 1 |
| Maldives | 0 |
| **Total (sum; some records tagged to multiple countries)** | 181 |

Records were screened for relevance to measles outbreak context, genotype or sequence data, molecular detection, phylogenetic analysis, vaccination context, or surveillance in Bangladesh or the broader South Asian region. The literature was used to contextualize public molecular evidence, not to estimate pooled effects. India accounted for 66.9% (111/166) of the regional molecular-surveillance literature in Search 1, paralleling the GenBank public-sequence-visibility asymmetry and reinforcing the regional literature-density pattern described in the main text.

**Post-search verification (June 16, 2026):** A no-date-filter re-query of the same three searches on June 16, 2026 returned 169 records for Search 1, 97 for Search 2, and 30 for Search 3, indicating that only 3 additional records for Search 1 and 4 for Search 2 had been indexed in PubMed after the May 21 search date (Search 3 yielded no new records). The 3 post-May-21 Search 1 records were: a Pakistan 2022–2023 outbreak study (Nawaz A, epub 2026-05-28; PMID 42208305), a Bangladesh resurgence commentary (Hossain S, 2026-05-27; PMID 42192469), and a news article on the 2026 Bangladesh outbreak (Anderer S, 2026-06-16; PMID 42172022). The fourth post-May-21 record unique to Search 2 was a Bangladesh SSPE case series (Alam S, epub 2026-06-05; PMID 42256338). None of these post-May-21 records changed the regional literature-density interpretation, and all operational numbers reported in the main text are from the May 21, 2026 search. The verification check is reported once here in the Appendix; the main-text Methods cross-references this single source.

### Appendix Section 2. GenBank/NCBI retrieval and curation

NCBI Nucleotide was queried on May 21, 2026, using `txid11234[Organism:exp]` combined with each of 9 regional country terms (the 8 South Asian Association for Regional Cooperation [SAARC] countries plus Myanmar). Records were retrieved as GenBank XML through NCBI E-utilities and de-duplicated by unique accession number. Country-term search hits describe the publicly visible landscape; geographic origin parsed from each unique record (`geo_loc_name` and standardized strain name) was used for genotype composition. Counts are indicators of public sequence visibility, not national incidence, prevalence, or reservoir size.

**Appendix Table 1. Country-term search hits and unique records, measles virus, 9 regional terms, May 21, 2026**

| Country term | Search hits |
|---|---|
| India | 2,345 |
| Pakistan | 1,645 |
| Nepal | 44 |
| Afghanistan | 42 |
| Bangladesh | 34 |
| Myanmar | 28 |
| Sri Lanka | 10 |
| Bhutan | 2 |
| Maldives | 1 |
| **Total hits (before de-duplication)** | **4,151** |
| **Unique accession-level records (after de-duplication)** | **4,109** |

Hits exceed unique records because some accessions are retrieved by more than one country term. Records lacking a parseable geographic origin (n = 131) are retained in the unique total but not assigned to a country of origin in Appendix Table 2.

### Appendix Section 3. Genotype composition of unique geolocated records

Genotypes were parsed from WHO-standardized bracket notation (e.g., [B3], [D8], [D9]) in strain names or feature annotations. Composition is reported among unique records geolocated to each country of origin.

**Appendix Table 2. Genotype composition by country of origin among 4,109 unique records**

| Country (origin) | Records | Genotype composition (n, %) |
|---|---|---|
| India | 2,227 | D8 1,903 (85.5%); D4 217 (9.7%); B3 73 (3.3%); D7 9 (0.4%); A 5 (0.2%); other/unlabelled 20 (0.9%) |
| Pakistan | 1,592 | B3 1,535 (96.4%); D4 56 (3.5%); H1 1 (0.1%) |
| Bangladesh | 32 | B3 19 (59.4%); D8 7 (21.9%); unlabelled 6 (18.8%) |
| Nepal | 18 | D8 8 (44.4%); D4 7 (38.9%); unlabelled 3 (16.7%) |
| Afghanistan | 40 | B3 33 (82.5%); H1 4 (10.0%); D4 3 (7.5%) |
| Myanmar | 28 | D9 27 (96.4%); D8 1 (3.6%) |
| Sri Lanka | 10 | D8 9 (90.0%); H1 1 (10.0%) |
| Bhutan | 2 | D8 2 (100.0%) |

All 32 Bangladesh-origin records have collection years between 2014 and 2019; none was collected after 2019. Across the entire post-2020 period the de-duplicated dataset contained 1,251 Pakistan-origin and 177 India-origin records but no Bangladesh-origin record.

### Appendix Section 4. N450 sequence comparison

The 2026 travel-associated complete genome PZ189094.1 (strain MVs/Queensland.AUS/02.26[B3]) was trimmed to its 450-nt carboxyl-terminal N-gene window (N450). It was compared with regional genotype B3 N450 records and with D8 and D9 outgroups. Two MAFFT alignments were produced, both using MAFFT `--auto`: (i) a 22-taxon alignment (19 Bangladesh B3 + 2 Meghalaya B3 + 1 Basirhat B3) used to generate main-text Figure 2, with percent identity calculated over 450 comparable columns; and (ii) a 12-taxon focused alignment (8 representative B3 + 2 outgroups D8 and D9 + 2 additional B3) used for Appendix Table 3, with percent identity calculated over 447 comparable columns. The 12-taxon subset is a representative subset used for table legibility and does not alter the central identity result: PZ189094.1 is 98.881% identical to its closest B3 comparators over 447 columns and 98.889% identical over 450 columns, and the small difference reflects only the number of comparable alignment columns and not biological difference. Gaps and ambiguous bases were excluded from pairwise percent identity in both alignments. Similarity was interpreted as evidence of relatedness within a regional B3 background only; it was not used to infer ancestry, origin, transmission direction, or local persistence. Genotypes were taken from submitter-supplied labels in the bracket notation (e.g., [B3], [D8], [D9]) rather than re-called against WHO reference strains (e.g., MVi/Illinois.USA/50.99[B3], GenBank AF266287) or WHO MeaNS; this is noted as a limitation in the main text.

**Appendix Table 3. Pairwise N450 identity of PZ189094.1 versus regional comparators (focused 12-taxon MAFFT alignment; 447 comparable columns)**

| Accession | Origin | Year | Genotype | N450 identity | Matches/ compared | nt differences |
|---|---|---|---|---|---|---|
| PZ106574.1 | Bajaur, Pakistan | 2026 | B3 | 99.553% | 445/447 | 2 |
| MW735989.1 | Basirhat, West Bengal, India | 2020 | B3 | 98.881% | 442/447 | 5 |
| MK628293.1 | Cox's Bazar, Bangladesh | 2017 | B3 | 98.881% | 442/447 | 5 |
| MK628292.1 | Bandarban, Bangladesh | 2016 | B3 | 98.881% | 442/447 | 5 |
| MK628286.1 | Bagerhat, Bangladesh | 2018 | B3 | 98.881% | 442/447 | 5 |
| MK183782.1 | Cox's Bazar, Bangladesh | 2017 | B3 | 98.881% | 442/447 | 5 |
| KX350060.1 | East Khasi Hills, Meghalaya, India | 2016 | B3 | 98.881% | 442/447 | 5 |
| KX350059.1 | East Khasi Hills, Meghalaya, India | 2016 | B3 | 98.881% | 442/447 | 5 |
| MK183787.1 | Comilla, Bangladesh | 2017 | B3 | 98.658% | 441/447 | 6 |
| JX905350.1 | Bago, Myanmar | 2010 | D9 (outgroup) | 90.604% | 405/447 | 42 |
| MK628291.1 | Mymensingh, Bangladesh | 2018 | D8 (outgroup) | 90.604% | 405/447 | 42 |

The single closest public N450 relative of PZ189094.1 is the contemporaneous 2026 Pakistan B3 record PZ106574.1 (2-nt difference), not a Bangladesh record. The historical Bangladesh and northeastern Indian (Meghalaya) B3 records are equally distant from PZ189094.1 (5-nt difference). In the primary 22-taxon MAFFT analysis underlying main-text Figure 2 (450-column window), PZ189094.1 was 98.889% identical (445/450; 5-nt difference) to the Meghalaya records, the Basirhat B3 record (MW735989.1), and 10 historical Bangladesh B3 records (KX671298.1, MK161512.1, MK161514.1, MK161516.1, MK183782.1, MK183783.1, MK183784.1, MK628286.1, MK628292.1, MK628293.1), with 9 further Bangladesh B3 records at 98.667% (444/450; 6-nt difference). The small difference between 98.881% and 98.889% reflects the number of comparable alignment columns (447 vs 450) and is not biologically meaningful. These data establish membership in a shared regional B3 background; they do not establish geographic origin or transmission direction.

#### Appendix Figure 1. Exploratory maximum-likelihood phylogeny (RAxML, GTR+GAMMA, 1,000 rapid bootstrap replicates)

The 13-taxon N450 alignment: the 2026 travel-associated genome PZ189094.1 (MVs/Queensland.AUS/02.26[B3]; N450 extracted in-frame from the complete genome), historical Bangladesh B3 (Bandarban 2016, Cox's Bazar 2017, Dhaka 2018, Comilla 2018, Bagerhat 2018), northeastern-Indian/border B3 (East Khasi Hills, Meghalaya 2016 ×2; Basirhat, West Bengal 2020), a contemporaneous 2026 Pakistan B3 (PZ106574.1, Bajaur, Khyber Pakhtunkhwa), Bangladesh D8 (Mymensingh 2018, MK628291.1) and India D8 (Basirhat, West Bengal 2020, MW672281.1), and a Myanmar D9 (Bago 2010, JX905350.1) as outgroup. Sequences were retrieved from GenBank, aligned with MAFFT (`--auto`), and subjected to maximum-likelihood search and 1,000 rapid bootstrap replicates. Branch-support values are reported at the inner nodes; inner-B3-node bootstrap values range from 6% to 84% (the B3 records differ by only a few nucleotides and the inner B3 substructure is not strongly resolved). The 2026 traveller groups with the 2026 Pakistan B3 at 97% bootstrap support and a tip-to-tip distance of ~0.0044 substitutions/site, while the 2014–2018 Bangladesh and the 2016 Meghalaya B3 records form a single tightly clustered B3 clade at 100% bootstrap support. The 2020 West Bengal Basirhat B3 (MW735989.1) is nested within the Bangladesh B3 cluster. The Bangladesh D8 and India D8 sequences form a separate clade with 100% bootstrap support, and the Myanmar D9 root (outgroup) connects at the base of the tree. Interpretation is limited: the N450 window is short and these B3 records differ by only a few nucleotides, so internal branching within the B3 clade is not strongly resolved; the phylogeny cannot attribute PZ189094.1 to a specific geographic source, establish transmission direction, or demonstrate local persistence. The figure is provided as an exploratory aid only and is not used to support any directional or ancestry claim in the main text.

**Appendix Figure 1.** Exploratory maximum-likelihood phylogeny of regional isolates

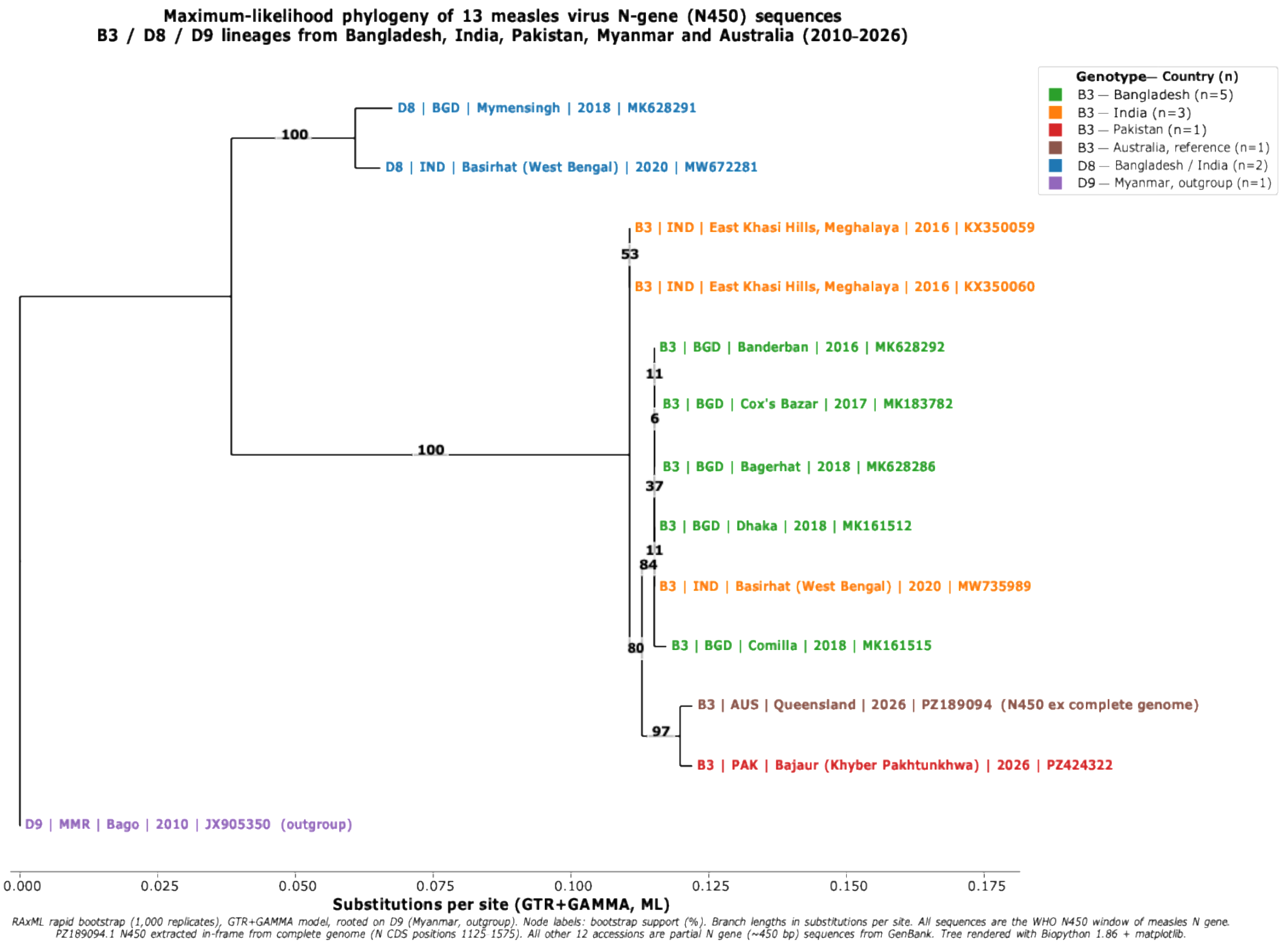


### Appendix Section 5. Immunization data and derived metrics

WHO/UNICEF Estimates of National Immunization Coverage (WUENIC, 2024 revision, released July 15, 2025) for the first and second doses of measles-containing vaccine (MCV1, MCV2), 2000–2024, were used as epidemiologic context. MCV1 series were also cross-checked against the WHO Global Health Observatory (indicator WHS8_110). For each of the 9 countries with WUENIC series (Afghanistan, Bangladesh, Bhutan, India, Maldives, Myanmar, Nepal, Pakistan, Sri Lanka), we derived the 2024 estimate, the 25-year mean and range, the pre-pandemic (2010–2019) and pandemic-era (2020–2024) means, and the MCV1–MCV2 gap; 2025 administrative coverage (WHO/UNICEF Joint Reporting Form, 2025 round) was compared with 2024 WUENIC. These values populate main-text Table 2. Coverage estimates were used as context and were not used to explain genotype distributions or GenBank record counts.

### Appendix Section 6. Reproducibility resources

All inputs and code are reproducible from the cited public sources. The submission package includes:

**Regional GenBank dataset:** de-duplicated 4,109-record accession-level table (CSV/XLSX) (05_data_saarc_myanmar_genbank_record.csv & 06_data_saarc_myanmar_genbank_record.xlsx) with accession, definition, organism, geographic origin, strain/isolate, collection date, parsed genotype, collection year, and transmission classification; plus a validation report documenting retrieval counts, de-duplication, and field completeness.

**Retrieval and curation script** (01_script_rebuild_measles_saarc_myanmar_genbank.py): NCBI E-utilities query, GenBank XML parsing, de-duplication, genotype/geography parsing, and validation.

**Sequence-comparison outputs:** input FASTA, MAFFT alignment, per-position variable-site report, and the pairwise N450 identity tables (Appendix Tables 3); plus the focused N450 input FASTA and Newick tree underlying Appendix Figure 1.

**Figure-generation scripts** (02_script_generate_charts.py, 03_script_generate_b3_tree_pro.py): main-text figures and the exploratory maximum-likelihood phylogeny (RAxML GTR+GAMMA, 1,000 rapid bootstrap replicates).

**Data and code availability.** All sequence records are available from NCBI GenBank under the accession numbers cited. WUENIC estimates are available from the WHO/UNICEF immunization data portal. The de-duplicated dataset, alignments, identity tables, and scripts are provided with this Appendix.
